# Making sense of non-randomized comparative treatment studies in times of Covid-19: A case study of tocilizumab

**DOI:** 10.1101/2021.04.06.21254612

**Authors:** Ruth Owen, Nawab Qizilbash, Sara Velázquez Díaz, José María Castellano Vázquez, Stuart Pocock

**Affiliations:** London School of Hygiene and Tropical Medicine, London, UK; Oxon Epidemiology, Madrid, Spain; Fundación de Investigación HM Hospitales, Grupo HM Hospitales, Madrid, Spain; Facultad de Medicina, Universidad CEU San Pablo, Madrid Spain; Centro Nacional de Investigaciones Cardiovasculares (CNIC), Instituto de Salud Carlos III, Madrid, Spain

## Abstract

**BACKGROUND:** Tocilizumab (TCZ) is an interleukin-6 inhibitor and the second established effective drug for the treatment of hospitalized patients with Covid-19. In this study, we sought to validate the recent positive findings from the randomised clinical trial RECOVERY and to evaluate the challenges in the analysis and interpretation of non-randomized comparative effectiveness studies in Covid-19.

**METHODS:** We performed a retrospective cohort study using an openly available database of hospitalised Covid-19 patients in Spain. The primary outcome was all-cause in-hospital mortality at 28 days. We used multivariable Fine and Gray competing risk models which adjusted for both fixed and time-variant confounders to investigate the effect of TCZ on the primary outcome.

**RESULTS:** We analysed 2547 patients hospitalised with Covid-19 between 1^st^ January and 28^th^ June 2020. Patients in the TCZ group tended to have more severe Covid-19 at admission, as measured by biomarkers of disease severity including CRP, D-dimer and LDH. At 28 days, 91 out of 440 TCZ patients had died compared to 267 out of 2107 patients in the control group. In multivariable analysis, there was no evidence of an association between TCZ and the primary outcome (adjusted hazard ratio 1.20, 95% CI 0.86 to 1.64, P=0.26).

**CONCLUSIONS:** Our observational study failed to find a benefit of TCZ on all-cause in-hospital mortality in Covid-19 patients compared with randomized trials, highlighting the impact that unmeasured confounding and other sources of bias can have in a retrospective observational setting. For future observational studies, we recommend prospective data collection to ensure all variables have the necessary quality, completeness and timing for reliable treatment evaluation.

## INTRODUCTION

Tocilizumab is the second established effective drug, after systematic corticosteroids, for the treatment of hospitalized patients with Covid-19. Tocilizumab is a monoclonal antibody that inhibits interleukin −6 (IL-6) and being repurposed from its use as an anti-inflammatory agent for rheumatoid arthritis and cytokine release syndrome to Covid-19 has not been straightforward.

IL-6 is cytokine released by macrophages in response to infection and stimulates inflammatory pathways. Levels of IL-6 are correlated with Covid-19 severity (1, 2).

In severe Covid-19 there is vascular inflammation and dysfunction (3, 4) and IL-6 promotes endothelial dysfunction and impairs vascular permeability. Tocilizumab inhibits this inflammatory process.

Given the lack of effective anti-viral agents to treat severe Covid-19, clinicians tried tocilizumab, among others, with early case reports indicating clinical and biochemical improvement (5-7). This was followed by reports of non-randomized comparative studies using retrospective data largely supporting the clinician-based impressions of benefit. This led to wider use of tocilizumab despite failure to show a survival benefit from early underpowered trials in severe Covid-19. The large RECOVERY trial has recently shown clear overall benefit (including reduced mortality) in hospitalized Covid-19 patients of all severities, in addition to systematic corticosteroids (8).

With knowledge of this background we conducted an analysis on a substantial individual patient database from multiple hospitals to evaluate the issues in planning analysis and interpretation of non-randomized comparative effectiveness studies in Covid-19. Besides the results themselves our goal is to provide guidance on appropriate methodology for other non-randomized comparative analyses of therapies in hospitalized patients with Covid-19 and help to understand the discrepancies between the randomized and non-randomized comparative studies of tocilizumab.

## METHODS

### Study setting and data source

We analysed data from the Covid Data Save Lives database, an anonymized dataset (9), which included electronic health records from 2547 patients hospitalized with COVID-19 in 17 hospitals within the Grupo HM Hospitales in Spain. This clinical dataset collects the different components of the COVID-19 treatment process, including detailed information on diagnoses, treatments, admissions, ICU admissions, vital signs, laboratory results, discharge and death. Data on medical history and co-morbidities were extracted from over 5,000 diagnostic and procedural records coded according to the international ICD-10 classification in its latest distributed version (10). Records of vital sign and laboratory analysis measurements were collected throughout admission, along with the date and time of collection. Information on tocilizumab (TCZ) administration was obtained from the medication table, including dose, number of doses and date of administration, and was identified by brand name and ATC5/ATC7 classification.

Patients were included if they had been admitted to any of the participating hospitals with a diagnosis of COVID POSITIVE or COVID PENDING between 1st January and 28th June 2020. Follow-up was from hospital admission until hospital discharge (either home or to another hospital/centre) or death. This study was approved by the ethics committee of HM Hospitales (approval number 20.05.1627-GHM).

### Study endpoints

The primary endpoint was all-cause in-hospital mortality at 28 days. Patients alive were censored at the earliest of end of study date (28^th^ June 2020), 28 days follow-up or transfer to another hospital.

### Statistical analysis

Patient characteristics at admission were summarized in the total cohort and compared in patients treated with TCZ versus those never treated with TCZ using Mann-Whitney U tests or independent t-tests for continuous variables and Fisher’s exact tests or chi-squared tests for categorical variables where appropriate.

Because patients discharged prior to 28 days were likely to have a substantially lower probability of death post discharge, we assumed that those discharged home were alive at 28 days by considering hospital discharge as a competing risk. The cumulative incidence function (CIF) of in-hospital mortality was plotted a) from time of admission in patients not treated with TCZ, that is patients in the control group and patients in the TCZ group censored at time of TCZ administration, and b) from time of TCZ administration in the TCZ group. Fine & Gray models (11) were used in univariable and multivariable analyses of the primary outcome to estimate subdistribution hazard ratios (HRs). sHRs have a similar interpretation to hazard ratios, except the risk set includes all patients discharged home earlier than 28 days from hospital admission. To account for immortal time bias, TCZ was modelled as a time-updated covariate. That is, patients would transition from ‘Not treated with TCZ’ to ‘Treated with TCZ’ at the time of TCZ administration and remain in the TCZ group until the end of follow-up.

Covariates considered in multivariable analysis were selected based on existing clinical knowledge and on risk factors of COVID-19 related mortality identified in previous studies (12-14) and are listed in **Supplementary Table 1** along with a summary of missing data. The distribution of each continuous variable was inspected and any variable that was highly positively skewed was log-transformed for analyses. Univariable analyses were first performed to explore the associations of patient characteristics at admission with all-cause mortality at 28 days in the dataset. A multivariable model was then fitted to investigate the effect of TCZ on 28-day mortality after adjustment for confounding. All potential confounders were first included in the model, then removed sequentially using a stepwise method with p>0.20 as the criterion for exclusion. Both age and sex were forced into the model.

In multivariable analyses, the following variables were fitted as time-updated covariates: saturated oxygen, diastolic blood pressure (DBP), systolic blood pressure (SBP), heart rate, alanine transaminase (ALT), aspartate transaminase (AST), C-reactive protein (CRP), creatinine, d-dimer, eosinophils, glucose, lactate dehydrogenase (LDH), lymphocytes, monocytes, neutrophils, platelet count, potassium, sodium, urea, white blood cell count (WBC) and treatment with steroids. To avoid adjusting for variables on the causal pathway of the association between TCZ and mortality, these covariates were only updated up to the point of TCZ administration for patients in the TCZ group. Additionally, the multivariable model was refitted using only the baseline values of the time-updated covariates, in order to explore the impact of ignoring time-variant confounding,

A subgroup analysis of CRP at admission was performed (CRP≥130mg/L vs CRP<130mg/L) by including an interaction term in the multivariable model.

Medical history and comorbidities were assumed to be absent if they had not been reported in the diagnostics table. The first vital sign or laboratory analysis measurement taken within 3 days of admission was considered the baseline value. For patients with no measurements reported during admission, baseline values were imputed using multiple imputation with chained equations and assumed constant throughout follow-up. The imputation models included the primary outcome, the Nelson-Aalen estimator of the cumulative hazard function and baseline variables with no missing data. Twenty imputation sets were generated and combined using Rubin’s rule (15).

Statistical analyses were performed using Stata v16.1 (StataCorp LP College Station, TX, USA). A two-sided p-value of 0.05 was considered statistically significant in all analyses.

## RESULTS

Of 2547 patients admitted to hospital with a diagnosis of COVID-19, 440 received at least one dose of TCZ. **Table 1** summarises patient characteristics at hospital admission in the total cohort and according to treatment with TCZ. Compared to the control group, patients receiving TCZ were younger (mean age 66.5 vs 68.2 years), had lower saturated oxygen (median 94.0% vs 95.3%), were more likely to be male (71.8% vs 56.1%) and obese (11.8% vs 6.9%). They also had higher AST, CRP, D-dimer and LDH levels, all of which are markers of disease severity in COVID-19 patients. In the control group, there was higher prevalence of dementia (4.7% vs 0.9%), renal disease (7.5% vs 4.1%) and diabetes (17.8% vs 12.3%) and controls tended to have a higher Charlson comorbidity index.

**Table 1.**
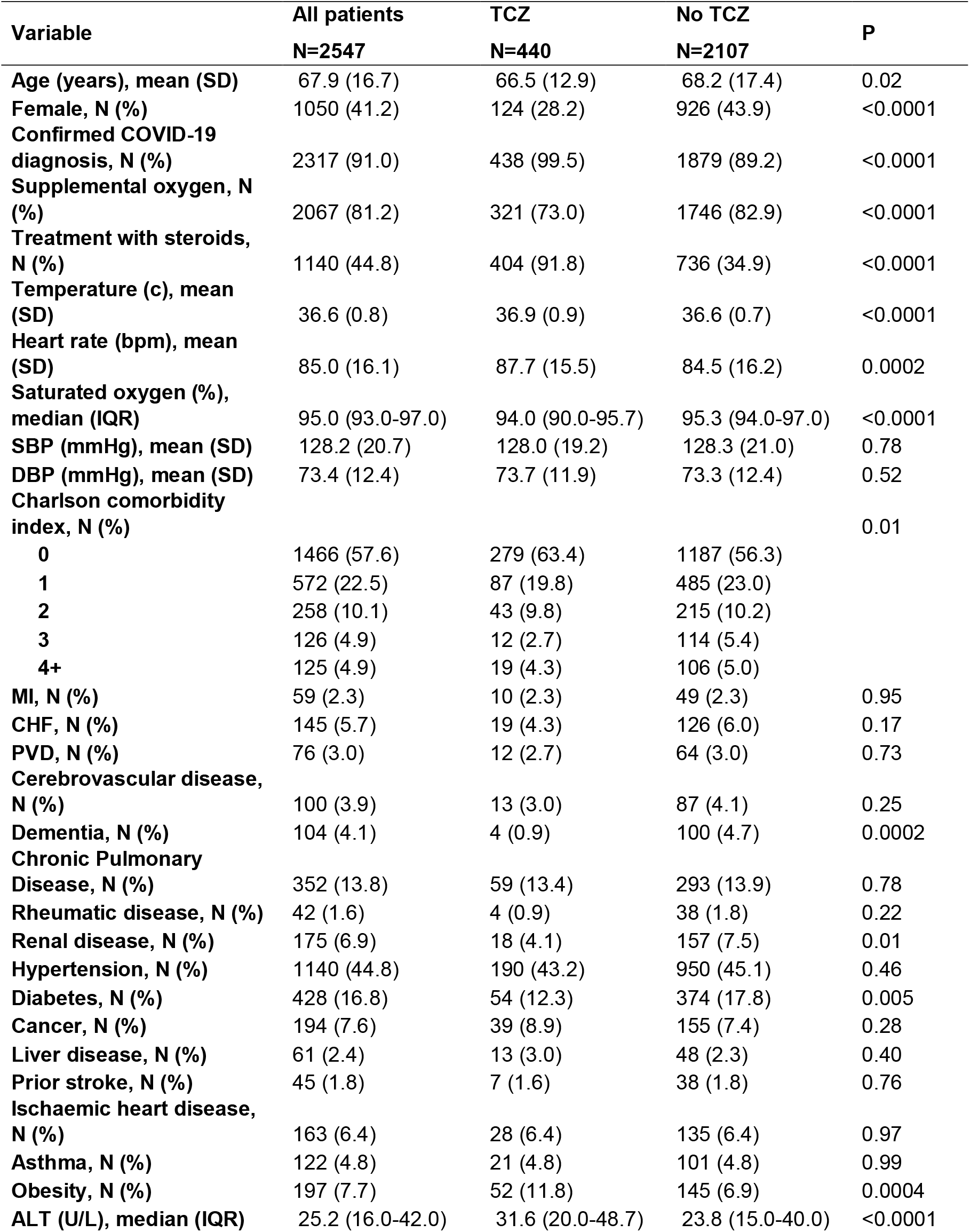
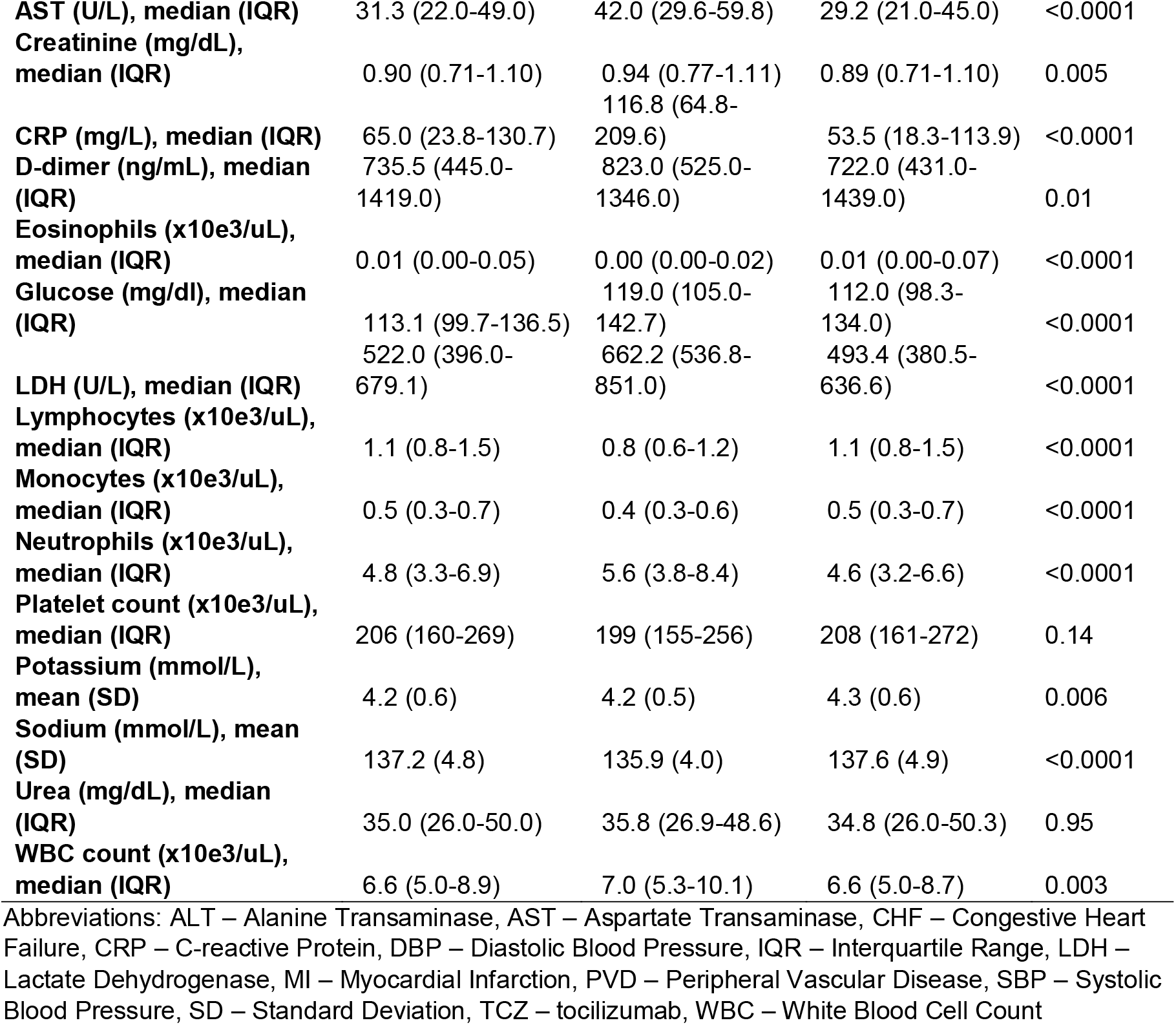
Patient characteristics at hospital admission

Patients in the control group were followed up for a median time of 6 days (IQR 4-9) from hospital admission. Patients in the TCZ group were followed up for a median time of 14 days (IQR 9-23) from hospital admission and a median time of 11 days (IQR 6-18) from TCZ administration. The median time to TCZ administration from hospital admission was 3 days (IQR 1-4) and the majority of TCZ patients received one dose (79.8%), with the remaining patients receiving two. At 28 days follow-up, a total of 358 patients (14.1%) had died, 1893 (74.3%) had been discharged home, 141 (5.5%) had been transferred to another hospital or centre and 155 (6.1%) patients remained in hospital.

The proportion of deaths at 28 days follow-up was higher in the TCZ group, 91 patients of whom died (20.7%) compared to the control group, 267 patients of whom died (12.7%).

**Figure 1A** shows the cumulative mortality from time of hospital admission in patients not treated with TCZ or up to the point of treatment, which is relatively constant for the first 9 days then appears to decline for the remainder of follow-up. However, this should be interpreted with caution as the censoring of patients at time of TCZ administration may lead to an over-optimistic estimate of survival due to the selection of more severe patients for treatment with TCZ. In contrast, there is a fairly steady increase in mortality from time of tocilizumab administration to 28 days follow-up in patients treated with tocilizumab (**Figure 1B**).

**Figure 1.**
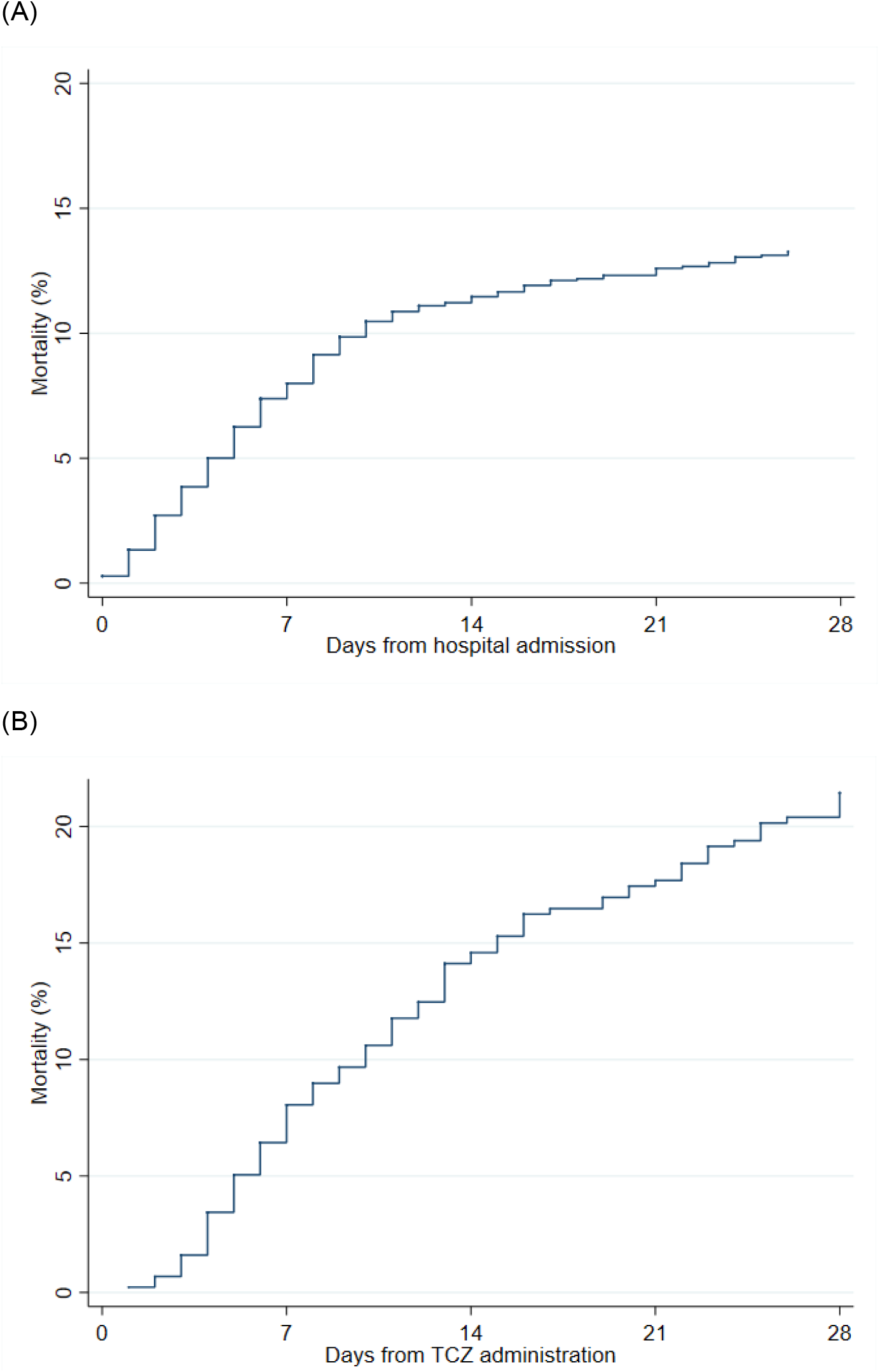
Cumulative incidence of mortality (A) from time of hospital admission in patients not treated with tocilizumab or up to the point of treatment and (B) from time of tocilizumab administration in 440 patients treated with tocilizumab. Abbreviations: TCZ - Tocilizumab

The univariable associations between each potential confounder and the primary endpoint are shown in **Supplementary Table 2**. In the unadjusted analysis, TCZ was associated with a higher risk of mortality (sHR 2.35, 95% CI 1.86 to 2.98, P<0.001, **Table 2**). Time-invariant confounders adjusted for in the multivariable model were age, sex, oxygen at admission, treatment with steroids, CHF, ischaemic heart disease, pulmonary disorder, cancer and PVD. Time-updated covariates were saturated oxygen, heart rate, ALT, AST, CRP, glucose, LDH, lymphocytes, monocytes, platelet count, sodium, urea and WBC. After adjustment for time-invariant confounding and baseline values of time-variant confounders, the harmful effect of TCZ decreased in magnitude but remained statistically significant (sHR 1.92, 95% CI (1.42, 2.60), p<0.001). After adjustment for time-variant and -invariant confounding the effect of TCZ was no longer statistically significant (sHR 1.20, 95% CI 0.86 to 1.64, P=0.26). Subgroup analysis showed some evidence of a difference in the effect of TCZ according to CRP at admission (P for interaction=0.03), suggesting a detrimental effect of TCZ in 1906 patients with a CRP<130 at hospital admission (sHR 1.66, 95% CI (1.11, 2.48)), whereas there swas no significant effect of TCZ in 641 patients with CRP≥130 (sHR 0.93, 95% CI (0.63, 1.37), **Supplementary Figure 1**).

**Table 2.** Unadjusted and adjusted effect of tocilizumab on 28-day all-cause mortality

|  | TCZ |  | No TCZ |  | Unadjusted |  | Adjusted for time invariant confounding<sup>2</sup> |  | Adjusted for time invariant and variant confounding<sup>3</sup> |  |
| --- | --- | --- | --- | --- | --- | --- | --- | --- | --- | --- |
|  | No. patients | No. deaths | No. patients | No. deaths | HR<sup>1</sup> (95% CI) | P | HR<sup>1</sup> (95% CI) | P | HR<sup>1</sup> (95% CI) | P |
| Mortality | 440 | 91 | 2107 | 267 | 2.35 (1.86, 2.98) | <0.001 | 1.92 (1.42, 2.60) | <0.001 | 1.20 (0.86, 1.64) | 0.26 |

Abbreviations: CI – Confidence interval, HR – Hazard Ratio, TCZ - Tocilizumab
<sup>1</sup>Hazard ratios were estimated using Fine & Gray models which considered discharge from hospital prior to 28 days as a competing risk
<sup>2</sup> Adjusted for age, sex, oxygen at admission, treatment with steroids, CHF, ischaemic heart disease, pulmonary disorder, cancer and PVD and for baseline values at hospital admission of saturated oxygen, heart rate, ALT, AST, CRP, glucose, LDH, lymphocytes, monocytes, platelet count, sodium, urea and WBC.
<sup>3</sup> Adjusted for age, sex, oxygen at admission, treatment with steroids, CHF, ischaemic heart disease, pulmonary disorder, cancer and PVD as time-invariant covariates and for saturated oxygen, heart rate, ALT, AST, CRP, glucose, LDH, lymphocytes, monocytes, platelet count, sodium, urea and WBC as time-varying covariates.

## DISCUSSION

Our non-randomized comparative analysis did not find the beneficial effect of TCZ established by the very recent large RECOVERY randomized trial. While the 95% confidence intervals of our analysis overlapped with that of RECOVERY, the point estimate did not fall within the 95% CI of the RECOVERY trial’s estimate. We found an apparent significant excess risk of death with TCZ in an unadjusted analysis but this attenuated to a much lesser non-significant excess after adjustment for many variables at baseline and further adjustment for time-varying covariates. The differences in the results from the fully adjusted multivariable model and the model with only adjustment for baseline values highlights the danger of ignoring time-variant confounding. However, the way to account for this type of confounding is not always clear and can require sophisticated statistical methods, resulting in effect estimates that are difficult to interpret. We took care to avoid adjustment of variables on the causal pathway of the association of interest. Allowance for missing data using multiple imputation made little difference. We did not find convincing evidence that the effect of TCZ was beneficial in patients with a high CRP as a marker of more severe disease. Allowing for the beneficial effect of corticosteroids made negligible difference.

Our findings also differ from three other non-randomized studies of TCZ at moderate risk of bias (16-18) which found more extreme beneficial effects than RECOVERY. Our findings are similar to the unadjusted analysis on this same but smaller and earlier dataset (19). We did not confirm the subgroup finding of benefit in patients with high CRP (19) which has also not been confirmed by other observational studies and the RECOVERY trial used CRP ≥ 75 mg/L as an inclusion criterion, with a median value of 143 (IQR: 107-207) in the randomized population. TCZ was administered to patients with severe Covid-19 based on the mortality rate of the group (and comparable to other observational studies and the severely ill subgroup in RECOVERY).

The reasons for these discrepancies are difficult to identify. Our population was reasonably large and included all, unselected hospitalized patients with moderate and severe Covid-19. RECOVERY found benefits in both these groups, dispelling earlier claims of the beneficial effects of TCZ only in severe or critically ill Covid-19 (20). The timing of administration of TCZ was soon after admission, without huge variation and all patients received mostly one or two doses on consecutive days, so unlikely to be factors affecting the results. Many covariates were included in our model comparable to other non-randomized studies and most established risk factors were confirmed in this dataset.

Our study has some limitations. Only death was analysed and other endpoints such as use of ICU and mechanical ventilation are important for hospital resource utilization in times of a pandemic. However, all-cause death is the outcome with least scope for misclassification and no missing data. We adjusted for competing risk. With sufficient data, if this endpoint cannot provide a valid finding other endpoints used in observational studies (16-18) are open to greater difficulties in analysis.

Other repurposed drugs which were used in the hospitals of the dataset were not included with the exception of corticosteroids but there is no good evidence for their beneficial effect (SOLIDARITY).

We did not use propensity scores for adjustment but our statistical models using multivariable adjustment were appropriate and there is little evidence that propensity score adjustments would give different results (21).

The quality of the data did not allow accurate analyses of the timing of administration of TCZ in relation to mechanical ventilation or date of ICU admission. Clinical severity was not always recorded or assessable at baseline or the time of administration of TCZ, however, the analysis used laboratory markers of severity such as CRP and D-dimer and LDH.

During the worst weeks of the pandemic from which this dataset arises, comorbidities at admission were not comprehensively collected, with some chronic conditions being recorded after admission. While many of the chronic conditions recorded after admission were ascribed to being present at baseline for analysis, the multivariable analysis of baseline variables may have been suboptimal, allowing for considerable residual confounding. Some key variables were absent such as smoking and BMI, and obesity defined clinically was present in only a minority of patients, again leading to potential residual confounding. Important time varying covariates were collected or recorded in a non-uniform, real-world fashion and this may have led to sub-optimal post-baseline adjustments. Time since symptom onset was also not recorded and there were a lot of missing data.

While negative controls for both exposure and endpoints have been proposed (22), we had a gold standard reference of a large reliable randomized trial with precise estimates containing large clinical subgroups for severity of covid-19 and use of corticosteroids. For the same reason, we did not calculate an E-value, a quantitative bias analysis to assess the strength of association between an unmeasured confounder and the exposure or outcome, conditional on measured covariates that would be necessary to fully explain observed effects (23).

In conclusion, our study failed to find a benefits of TCZ on all-cause in-hospital mortality in Covid-19 compared with randomized trials, which we believe is due to a combination of residual confounding and suboptimal adjustment with time varying covariates, largely due to deficiencies in the quality of data. Other non-randomized comparative studies at moderate risk of bias found exaggerated benefits compared with randomized trials. Thus, many non-randomized comparative studies of treatments using retrospective data appear unreliable in hospitalised Covid-19. For future observational studies, we recommend prospective data collection to ensure all variables have the necessary quality, completeness and timing for reliable treatment evaluation.

## Data Availability

Covid Data Saves Lives is an anonymous dataset made freely available to the international medical and scientific community by HM Hospitales

https://www.hmhospitales.com/coronavirus/covid-data-save-lives/english-version

