## Supplementary Appendix for "Making sense of non-randomized comparative treatment studies in times of Covid-19: A case study of tocilizumab"

**Supplementary Table 1.** Potential confounders considered in the multivariable analysis and missing data

| Variable | No. patients with missing values at admission (%) | No. patients with missing values during total FU (%) |
|---|---|---|
| **Age (years)** | 0 (0.0) | |
| **Sex** | 0 (0.0) | |
| **Confirmed COVID-19 diagnosis** | 0 (0.0) | |
| **Supplemental oxygen** | 0 (0.0) | |
| **Treatment with steroids** | 0 (0.0) | |
| **Temperature (c)*** | 65 (2.6) | 38 (1.5) |
| **Heart rate (bpm)*** | 80 (3.1) | 47 (1.8) |
| **Saturated oxygen (%)*** | 74 (2.9) | 43 (1.7) |
| **SBP (mmHg)*** | 250 (9.8) | 191 (7.5) |
| **DBP (mmHg)*** | 251 (9.9) | 192 (7.5) |
| **Charlson comorbidity index** | 0 (0.0) | |
| **Prior MI** | 0 (0.0) | |
| **CHF** | 0 (0.0) | |
| **PVD** | 0 (0.0) | |
| **Cerebrovascular disease** | 0 (0.0) | |
| **Dementia** | 0 (0.0) | |
| **Pulmonary Disease** | 0 (0.0) | |
| **Renal disease** | 0 (0.0) | |
| **Hypertension** | 0 (0.0) | |
| **Diabetes** | 0 (0.0) | |
| **Cancer** | 0 (0.0) | |
| **Liver disease** | 0 (0.0) | |
| **Prior stroke** | 0 (0.0) | |
| **Ischemic heart disease** | 0 (0.0) | |
| **Obesity** | 0 (0.0) | |
| **ALT (U/L)*** | 350 (13.7) | 247 (9.7) |
| **AST (U/L)*** | 340 (13.3) | 243 (9.5) |
| **Creatinine (mg/dL)*** | 250 (9.8) | 187 (7.3) |
| **CRP (mg/L)*** | 250 (9.8) | 186 (7.3) |
| **D-dimer (ng/mL)*** | 551 (21.6) | 378 (14.8) |
| **Eosinophils (x10e3/uL)*** | 230 (9.0) | 171 (6.7) |
| **Glucose (mg/dl)*** | 280 (11.0) | 209 (8.2) |
| **LDH (U/L)*** | 302 (11.9) | 225 (8.8) |
| **Lymphocytes (x10e3/uL)*** | 212 (8.3) | 156 (6.1) |
| **Monocytes (x10e3/uL)*** | 230 (9.0) | 171 (6.7) |
| **Neutrophils (x10e3/uL)*** | 212 (8.3) | 156 (6.1) |
| **Platelet count (x10e3/uL)*** | 212 (8.3) | 156 (6.1) |

| | | |
|---|---|---|
| **Potassium (mmol/L)*** | 278 (10.9) | 206 (8.1) |
| **Sodium (mmol/L)*** | 277 (10.9) | 204 (8.0) |
| **Urea (mg/dL)*** | 291 (11.4) | 225 (8.8) |
| **WBC count (x10e3/uL)*** | 212 (8.3) | 156 (6.1) |

Abbreviations: ALT – Alanine Transaminase, AST – Aspartate Transaminase, CHF – Congestive Heart Failure, CRP – C-reactive Protein, DBP – Diastolic Blood Pressure, IQR – Interquartile Range, LDH – Lactate Dehydrogenase, MI – Myocardial Infarction, PVD – Peripheral Vascular Disease, SBP – Systolic Blood Pressure, WBC – White Blood Cell Count

* Modelled as time-updated covariates in multivariable analyses

**Supplementary Table 2.** Univariable associations of patient characteristics at admission with 28-day mortality

| Variable | No. deaths | HR[1] (95% CI) | P |
|---|---|---|---|
| **Age (years)** | | 1.08 (1.07, 1.09) | <0.0001 |
| **Female** | 130 | 0.73 (0.59, 0.91) | 0.005 |
| **Confirmed COVID-19 diagnosis** | 347 | 0.94 (0.65, 1.35) | 0.72 |
| **Supplemental oxygen** | 304 | 1.09 (0.83, 1.42) | 0.53 |
| **Treatment with steroids** | 247 | 2.06 (1.67, 2.56) | <0.0001 |
| **Temperature (c)** | | 0.92 (0.81, 1.05) | 0.21 |
| **Heart rate (bpm)[2]** | | 1.05 (0.98, 1.12) | 0.17 |
| **Saturated oxygen (%)** | | 0.95 (0.94, 0.97) | <0.0001 |
| **SBP (mmHg)[2]** | | 0.99 (0.94, 1.04) | 0.67 |
| **DBP (mmHg)[2]** | | 0.85 (0.78, 0.92) | 0.0001 |
| **MI** | 18 | 2.44 (1.54, 3.88) | 0.0001 |
| **CHF** | 41 | 2.26 (1.62, 3.17) | <0.0001 |
| **PVD** | 17 | 1.71 (1.06, 2.77) | 0.03 |
| **Cerebrovascular disease** | 33 | 2.62 (1.83, 3.74) | <0.0001 |
| **Dementia** | 35 | 3.08 (2.19, 4.34) | <0.0001 |
| **Pulmonary Disease** | 74 | 1.66 (1.29, 2.15) | <0.0001 |
| **Rheumatic disease** | 5 | 0.83 (0.34, 2.03) | 0.69 |
| **Renal disease** | 51 | 2.46 (1.84, 3.29) | <0.0001 |
| **Hypertension** | 235 | 2.12 (1.71, 2.62) | <0.0001 |
| **Diabetes** | 86 | 1.55 (1.22, 1.98) | 0.0004 |
| **Cancer** | 60 | 2.46 (1.85, 3.26) | <0.0001 |
| **Liver disease** | 8 | 0.89 (0.45, 1.78) | 0.74 |
| **Prior stroke** | 14 | 2.42 (1.45, 4.05) | 0.0007 |
| **Ischemic heart disease** | 40 | 1.89 (1.36, 2.61) | 0.0001 |
| **Asthma** | 15 | 0.79 (0.46, 1.34) | 0.38 |
| **Obesity** | 25 | 0.81 (0.53, 1.23) | 0.32 |
| **Charlson comorbidity index** | | 1.25 (1.20, 1.29) | <0.0001 |
| **ALT (U/L)[3]** | | 0.71 (0.61, 0.83) | <0.0001 |
| **AST (U/L)[3]** | | 1.53 (1.34, 1.76) | <0.0001 |
| **Creatinine (mg/dL)[3]** | | 4.54 (3.50, 5.89) | <0.0001 |
| **C-reactive protein (mg/L)[3]** | | 2.08 (1.86, 2.32) | <0.0001 |
| **D-dimer (ng/mL)[3]** | | 1.51 (1.41, 1.62) | <0.0001 |
| **Eosinophils (x10e3/uL)[3]** | | 0.84 (0.79, 0.88) | <0.0001 |
| **Glucose (mg/dl)[3]** | | 4.12 (3.08, 5.50) | <0.0001 |
| **LDH (U/L)[3]** | | 4.13 (3.46, 4.94) | <0.0001 |
| **Lymphocytes (x10e3/uL)[3]** | | 0.49 (0.41, 0.59) | <0.0001 |
| **Monocytes (x10e3/uL)[3]** | | 0.94 (0.76, 1.15) | 0.53 |
| **Neutrophils (x10e3/uL)** | | 1.12 (1.10, 1.14) | <0.0001 |
| **Platelet count (x10e3/uL)[3]** | | 0.56 (0.44, 0.71) | <0.0001 |
| **Potassium (mmol/L)** | | 1.47 (1.20, 1.81) | 0.0002 |

| | | |
|---|---|---|
| **Sodium (mmol/L)** | 1.07 (1.05, 1.10) | <0.0001 |
| **Urea (mg/dL)[3]** | 4.84 (4.18, 5.59) | <0.0001 |
| **WBC count (x10e3/uL)[3]** | 3.17 (2.47, 4.06) | <0.0001 |

Abbreviations: ALT – Alanine Transaminase, AST – Aspartate Transaminase, CHF – Congestive Heart Failure, CRP – C-reactive Protein, DBP – Diastolic Blood Pressure, HR – Hazard Ratio, IQR – Interquartile Range, LDH – Lactate Dehydrogenase, MI – Myocardial Infarction, PVD – Peripheral Vascular Disease, SBP – Systolic Blood Pressure, WBC – White Blood Cell Count

[1] Hazard ratios were estimated using Fine & Gray models which considered discharge from hospital prior to 28 days as a competing risk

[2] Per 10 units

[3] Log transformed

**Supplemental Figure 1.** Subgroup analysis of mortality at 28 days by C-reactive protein (CRP) level at admission (CRP<130mg/L vs CRP≥130mg/L).

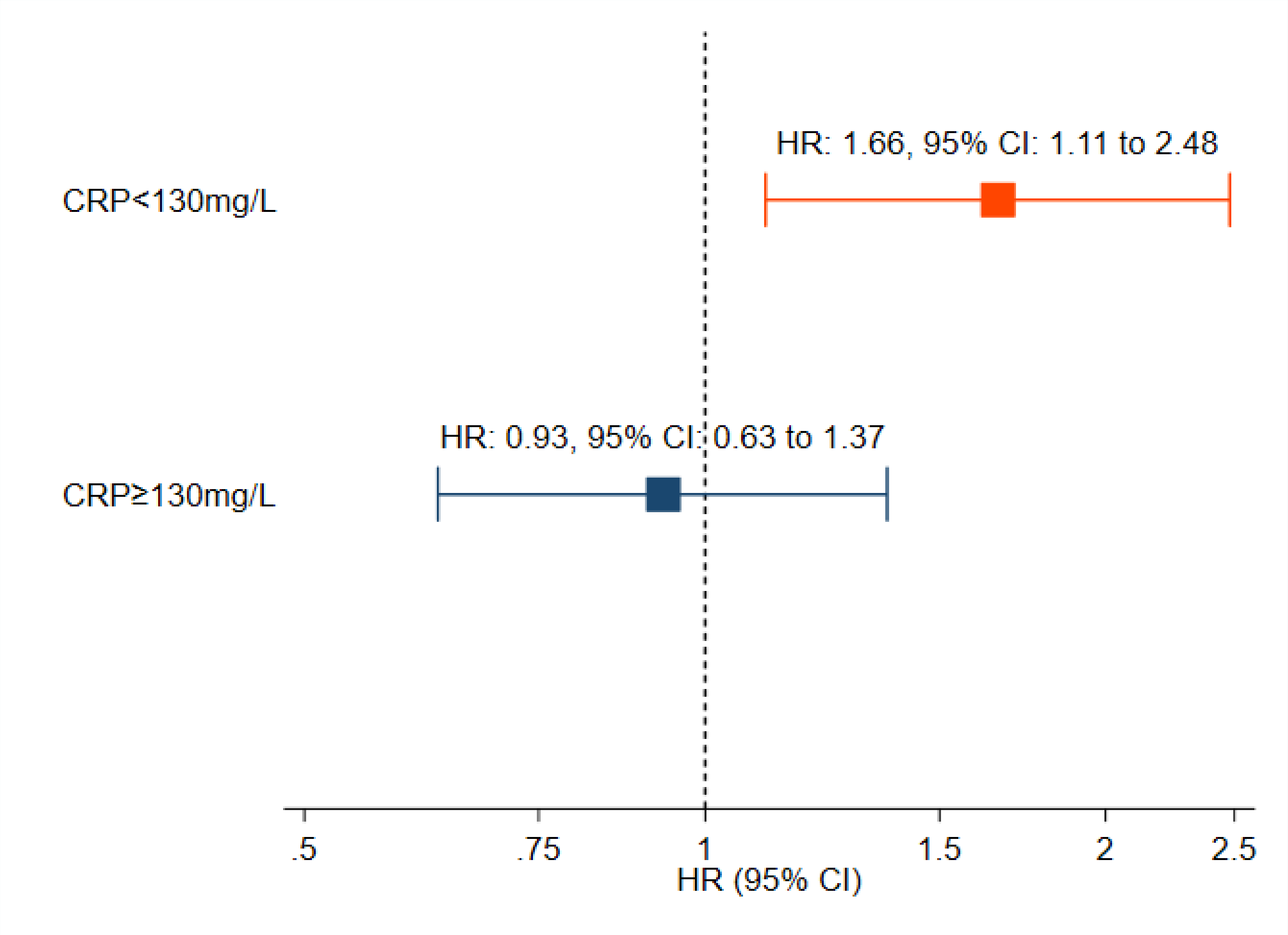

Abbreviations: CI – Confidence Interval, CRP – C-Reactive Protein, HR – Hazard Ratio
Hazard ratios were estimated using Fine & Gray models which considered discharge from hospital prior to 28 days as a competing risk with an interaction term between tocilizumab and CRP≥130mg/L at baseline and adjusted for age, sex, oxygen at admission, treatment with steroids, CHF, ischaemic heart disease, pulmonary disorder, cancer and PVD and for baseline values at hospital admission of saturated oxygen, heart rate, ALT, AST, glucose, LDH, lymphocytes, monocytes, platelet count, sodium, urea and WBC as time-varying covariates.
